# Analytical validation and amyloid-status discrimination of a high-throughput, research-use-only plasma p-Tau217 immunoassay

**DOI:** 10.64898/2026.08.31.26361836

**Authors:** Paul Wynveen, Andrew Becker, Scott Levin, Ben Dumke, Nathan Hoekstra, Katharine Hoffmann, Chris Knutson, Justin Lengfeld, Peng Li, Jeff Radcliff, Kinal Bhatt, Henrik Zetterberg, Andrea Lessa Benedet, Mark Holland, Corey M. Carlson, Jeremiah S. Hinson

**Author notes:** Drs. Wynveen and Becker contributed equally (co-first authors). Drs. Holland, Carlson, and Hinson contributed equally (co-senior authors). Corresponding Author*: Jeremiah Hinson, MD, PhD, Danaher Diagnostics, 2200 Pennsylvania Ave, NW Suite 800W, Washington, DC, USA.

## Abstract

**Background:** Plasma phosphorylated tau at threonine 217 (p-Tau217) is a leading blood-based biomarker for Alzheimer’s disease (AD). Robust analytical characterization on high-throughput platforms is essential for research use and clinical translation.

**Objective:** To evaluate the analytical performance of an automated plasma p-Tau217 immunoassay and characterize its discrimination of PET-defined amyloid status.

**Methods:** We performed analytical validation of the Access Research Use Only (RUO) plasma p-Tau217 immunoassay on the Beckman Coulter DxI 9000 Access Immunoassay Analyzer and evaluated biomarker discrimination of PET-defined amyloid pathology in a subset of the Bio-Hermes-001 cohort spanning the symptomatic cognitive continuum (mild cognitive impairment or mild AD dementia; cognitively unimpaired participants excluded; n = 449). Analytical precision, sensitivity, linearity, specificity, interference, and sample stability were assessed per Clinical and Laboratory Standards Institute guidelines. Discrimination of PET-defined amyloid status was evaluated using receiver operating characteristic curve and indeterminate zone analyses.

**Results:** The assay demonstrated high precision (within-laboratory CV ≤7.1%), excellent sensitivity (limit of detection 0.018–0.021 pg/mL), linearity across the analytical measuring range (R² > 0.99), strong epitope specificity (≤1.0% cross-reactivity with other tau phosphoisoforms), and minimal interference from over 60 endogenous and exogenous substances. In 449 research participants plasma p-Tau217 showed strong discrimination between amyloid-positive and amyloid-negative groups (AUC 0.881; 95% CI 0.846–0.915). Application of indeterminate zones systematically improved classification metrics at the cost of fewer definitive classifications.

**Conclusions:** These findings support the Access p-Tau217 (RUO) assay as a robust, high-throughput assay for plasma biomarker-based discrimination of PET-defined amyloid pathology in AD applications.

## Introduction

Alzheimer’s disease (AD), the leading cause of dementia worldwide, is characterized biologically by the accumulation of extracellular β-amyloid plaques and intracellular neurofibrillary tangles composed of hyperphosphorylated tau.^1,2^ Accurate characterization of these pathological processes is central to research efforts aimed at understanding disease mechanisms, staging disease along the AD continuum, and evaluating emerging therapeutic strategies.^3–5^ Cerebrospinal fluid (CSF) biomarkers and positron emission tomography (PET) imaging can detect AD-related pathology with high accuracy, but their widespread use is limited by invasiveness, cost and accessibility.^6–9^ These constraints have driven substantial interest in blood-based biomarkers as scalable, minimally invasive tools for AD research and clinical application. Robust analytical characterization of high-throughput assay platforms is essential to support both the research pipeline and translational pathway underlying these efforts.

Phosphorylated tau at threonine 217 (p-Tau217) has emerged as one of the most informative plasma biomarkers associated with AD pathology. Prior studies have demonstrated that plasma p-Tau217 distinguishes AD from other neurodegenerative conditions and correlates strongly with established markers of amyloid and tau pathology measured by CSF and PET.^10,11^ Plasma p-Tau217 levels increase early along the AD continuum, often in parallel with amyloid abnormalities, supporting its utility for biological characterization of disease processes in observational and translational research settings.^12–14^ Notably, several plasma p-Tau217 assays have received regulatory clearance in Europe,^6^ and a plasma p-Tau217/Aβ42 ratio assay has been cleared by the U.S. Food and Drug Administration,^15^ heightening the need for rigorous analytical characterization of the high-throughput platforms on which such measurements are performed.

In this study, we describe the analytical performance characteristics of the plasma Access p-Tau217 research use only (RUO) immunoassay implemented on the high-throughput DxI 9000 Access Immunoassay Analyzer (Beckman Coulter, Brea, CA) and evaluate its ability to discriminate between samples from individuals with PET-defined amyloid positivity and controls. We report assay precision, sensitivity, linearity, analytical specificity, and sample stability, and provide a detailed assessment of assay performance across a diverse research cohort, including a systematic evaluation of indeterminate zone strategies relevant to biomarker implementation.

## Methods

### Access p-Tau217 (RUO) assay

All p-Tau217 assay testing was performed on the DxI 9000 Access Immunoassay Analyzer (Beckman Coulter; Brea, CA), a fully automated chemiluminescent immunoassay platform designed for high-throughput laboratory testing. The system employs paramagnetic particle-based immunoassays with direct alkaline phosphatase-mediated chemiluminescent detection. The instrument aspirates samples directly from primary tubes or system sample cups and performs all assay steps, including reagent handling, incubation, washing, detection, and data processing. Details of the instrument and its operating principles have been described previously.^16–18^

The Access p-Tau217 (RUO) assay is a two-step sandwich immunoenzymatic assay. In the first step, K2EDTA plasma sample, reaction buffer, and monoclonal anti-p-Tau217 antibody-coated paramagnetic particles are incubated and washed. In the second step, an alkaline phosphatase-conjugated anti-tau monoclonal antibody is added, followed by incubation, wash, and addition of a chemiluminescent substrate. The emitted light is directly proportional to the concentration of p-Tau217 in the sample.

### Analytical validation

We performed a comprehensive analytical characterization of the Access p-Tau217 (RUO) immunoassay. Validation studies were conducted in accordance with Clinical and Laboratory Standards Institute (CLSI) guidelines, including EP05-A3 (precision),^19^ EP06-A (linearity),^20^ EP07 (interference),^21^ and EP17-A2 (detection capability).^22,23^ Plasma specimens used for analytical characterization (precision, analytical sensitivity, linearity, and analytical specificity) were obtained from commercial biospecimen vendors, and specimens used for the sample-stability study were provided by the University of Gothenburg. Specimen sources for each analytical study are detailed in the Supplement.

#### Precision

Assay precision was evaluated for six native K2EDTA plasma samples containing endogenous p-Tau217 levels spanning the analytical measurement range (AMR) and three quality controls. Each sample was tested in duplicate across forty runs over eight days using a single reagent lot and single analyzer. Within-run, between-run, between-day, and within laboratory standard deviation (SD) and coefficients of variation (CV%) were calculated.

#### Analytical sensitivity

Limit of blank (LoB), limit of detection (LoD), and limit of quantitation (LoQ) were estimated using two reagent lots and two analyzers.^22^ To estimate LoB, four blank samples were tested in replicates of five across three days. LoB was determined using a non-parametric method as the 95^th^ percentile of replicates for each reagent lot and instrument combination. LoD was estimated using five low-concentration p-Tau217 plasma samples tested with five replicates per run across two runs daily for five days. LoD was calculated as the LoB plus the low-concentration within-lab standard deviation multiplied by the 95^th^ percentile of the standard normal distribution. For LoQ, 12 low-concentration p-Tau217 samples were tested in replicates of five across two runs daily over five days. A variance components model estimated within-laboratory percent coefficient of variation (%CV) for each sample, instrument, and reagent lot combination. A log-log quadratic precision profile model was fitted to within-laboratory %CV versus observed mean, and the fitted profile determined the 20% CV LoQ.

#### Linearity

Linearity was evaluated using plasma samples spanning the assay’s AMR.^20^ A native low-concentration sample and a high-concentration sample (native plasma supplemented with recombinant p-Tau217 antigen) were prepared. Seven intermediate concentrations were created by adding incrementally larger proportions of high concentration sample diluted with the low sample covering the full AMR. The low sample was tested in eight replicates per run, and all others were tested in four replicates per run using one reagent lot on a single analyzer. In addition to the full-range linearity study, two additional linearity panels covering the expected physiological concentration range were evaluated using native samples for low and high concentrations with eight intermediate mixtures, tested across two reagent lots. Data were analyzed using weighted linear regression of observed versus expected results.

#### Analytical specificity

Assay specificity was assessed by testing endogenous and exogenous interferents at p-Tau217 concentrations of approximately 0.2 pg/mL and 0.7 pg/mL. Test samples with potential interferent were compared to control samples. Proportional amounts of solvent were added to control samples and compared to the test samples spiked with solvent-dissolved interferents. Interference was reported as the percentage difference between test and control samples.

Additionally, cross-reactivity was evaluated by testing various phosphorylation sites on tau at p-Tau217 concentrations of approximately 0.2 pg/mL and 0.7 pg/mL. Test samples with potential cross-reactants were compared to control samples. Proportional amounts of solvent were added to control samples and compared to test samples spiked with solvent-dissolved cross-reactants. Cross-reactivity was reported as the percentage difference between test and control samples. Analytical specificity studies were performed using a single analyzer and one reagent lot, with at least five replicates per sample.

#### Sample stability

Sample stability was evaluated on five plasma samples under three storage and handling conditions; samples were tested at baseline, stored at room temperature (20 – 25°C) between 6 and 24 hours, and stored refrigerated (2-8 °C) between 24 and 48 hours. An additional set of five samples were tested across 1 to 6 freeze-thaw cycles in addition to being stored for -20 °C for 30 days. Tests for each condition were performed in singlicate, and the percentage difference from baseline results was evaluated.

### Discrimination of PET-defined biomarker groups

#### Sample source

To evaluate performance of the Access (RUO) p-Tau217 assay for detecting PET-defined amyloid pathology, we analyzed plasma samples from 449 participants in the Bio-Hermes-001 study; a US-based multicenter study of over 1,000 participants spanning the cognitive continuum (cognitively unimpaired, mild cognitive impairment, and mild AD). For the present analysis, participants originally enrolled in the cognitively unimpaired sub-cohort were excluded to better reflect the population most likely to benefit from this assay. The resulting sample therefore comprised participants with mild cognitive impairment or mild AD dementia—a cognitively impaired, symptomatic population.^24,25^

Participants were classified as amyloid-negative or amyloid-positive by central visual read of florbetapir F-18 (Amyvid) PET; reads were performed by trained specialists (IXICO, London, UK) following the manufacturer’s read process, with the standardized uptake value ratio (SUVR) available to the reader.^24,25^ Available clinical data included age, sex, race, ethnicity, Mini-Mental State Examination (MMSE) scores, and diagnosis of probable AD. Probable AD was defined by an NIA-AA diagnosis of probable AD^3^ verified through medical records or, alternatively, by predefined screening criteria indicating cognitive impairment and functional decline, as detailed in the Bio-Hermes-001 study.^24^

Plasma was collected and processed using standardized cohort protocols and shipped to Beckman Coulter on dry ice for blinded p-Tau217 analysis. All participants provided written informed consent under protocols approved by the relevant institutional review boards.

#### Measurement of p-Tau217

Plasma p-Tau217 concentrations were measured using the Access p-Tau217 (RUO) assay and performed on the DxI 9000 Access Immunoassay Analyzer. All samples were tested in accordance with manufacturer protocols, with automated incubation, washing, detection, and result calculation. Laboratory personnel were blinded to clinical information during testing.

#### Statistical assessment of biomarker group discrimination

Receiver operating characteristic (ROC) analyses were performed to quantify the ability of plasma p-Tau217 concentrations to discriminate between PET-defined biomarker groups (amyloid negative or positive). Classification metrics, including sensitivity, specificity, positive predictive value (PPV), negative predictive value (NPV), and positive and negative likelihood ratios, were calculated across a range of prespecified concentration thresholds to describe threshold-dependent behavior of the assay in this research setting. Thresholds and classification metrics were evaluated for research characterization purposes only and are not intended to define clinical decision limits.

To explore how expanding ranges of overlapping biomarker concentrations affect classification characteristics, indeterminate zones ranging from 0% to 30% of the cohort distribution were applied, generating paired lower and upper concentration thresholds for each scenario. All metrics were calculated with respect to PET derived amyloid status and are reported as statistical descriptors of biomarker group separation within this observational cohort. Confidence intervals were calculated using the Wilson interval or, for likelihood ratios, the method of Simel et al.^26,27^ All analyses were performed using R version 4.1.3 (epiR package), JMP version 16, and Analyze-It version 5.92.

## Results

### Analytical validity

#### Precision

Representative precision data from one reagent lot are shown in Table 1. High precision was demonstrated across all concentrations (mean pg/mL 0.111 - 2.159), with maximum %CVs of 6.6% within run (repeatability), 2.4% between run, 2.0% between day, and 7.1% within laboratory.

**Table 1.** Analytic precision.

| Sample | N | Mean<br>(pg/mL) | Within Run |  | Between Run |  | Between Day |  | Within<br>Laboratory |  |
| --- | --- | --- | --- | --- | --- | --- | --- | --- | --- | --- |
|  |  |  | SD | %CV | SD | %CV | SD | %CV | SD | %CV |
| 1 | 80 | 0.111 | 0.007 | 6.4% | 0.003 | 2.4% | 0.002 | 1.7% | 0.008 | 7.1% |
| 2 | 80 | 0.129 | 0.008 | 6.6% | 0.000 | 0.0% | 0.000 | 0.2% | 0.009 | 6.6% |
| 3 | 80 | 0.158 | 0.008 | 5.3% | 0.003 | 1.6% | 0.003 | 1.6% | 0.009 | 5.7% |
| 4 | 80 | 0.268 | 0.017 | 6.2% | 0.000 | 0.0% | 0.005 | 2.0% | 0.018 | 6.5% |
| 5 | 80 | 0.388 | 0.014 | 3.7% | 0.005 | 1.4% | 0.004 | 1.0% | 0.016 | 4.1% |
| 6 | 80 | 0.457 | 0.018 | 4.0% | 0.005 | 1.1% | 0.008 | 1.8% | 0.021 | 4.5% |
| 7 | 80 | 0.737 | 0.022 | 2.9% | 0.008 | 1.0% | 0.011 | 1.4% | 0.025 | 3.4% |
| 8 | 80 | 1.135 | 0.046 | 4.0% | 0.000 | 0.0% | 0.013 | 1.2% | 0.047 | 4.2% |
| 9 | 80 | 2.159 | 0.076 | 3.5% | 0.019 | 0.9% | 0.013 | 0.6% | 0.079 | 3.7% |

Precision was maintained at the lowest concentrations evaluated, confirming reliable performance of the assay near its lower measurement limits.

#### Analytical sensitivity

The p-Tau217 assay demonstrated high analytical sensitivity. The LoB was estimated as 0.011 pg/mL for both reagent lots. Across two reagent lots, the estimated LoDs were 0.021 pg/mL (95% CI 0.020 - 0.023 pg/mL) and 0.018 pg/mL (95% CI 0.017 - 0.019 pg/mL). The assay’s LoQ precision profile is shown in Figure 1; the estimated LoQs across two reagent lots were 0.032 pg/mL (95% CI 0.021 - 0.036 pg/mL) and 0.018 pg/mL (95% CI 0.014 - 0.023 pg/mL).

**Figure 1.**
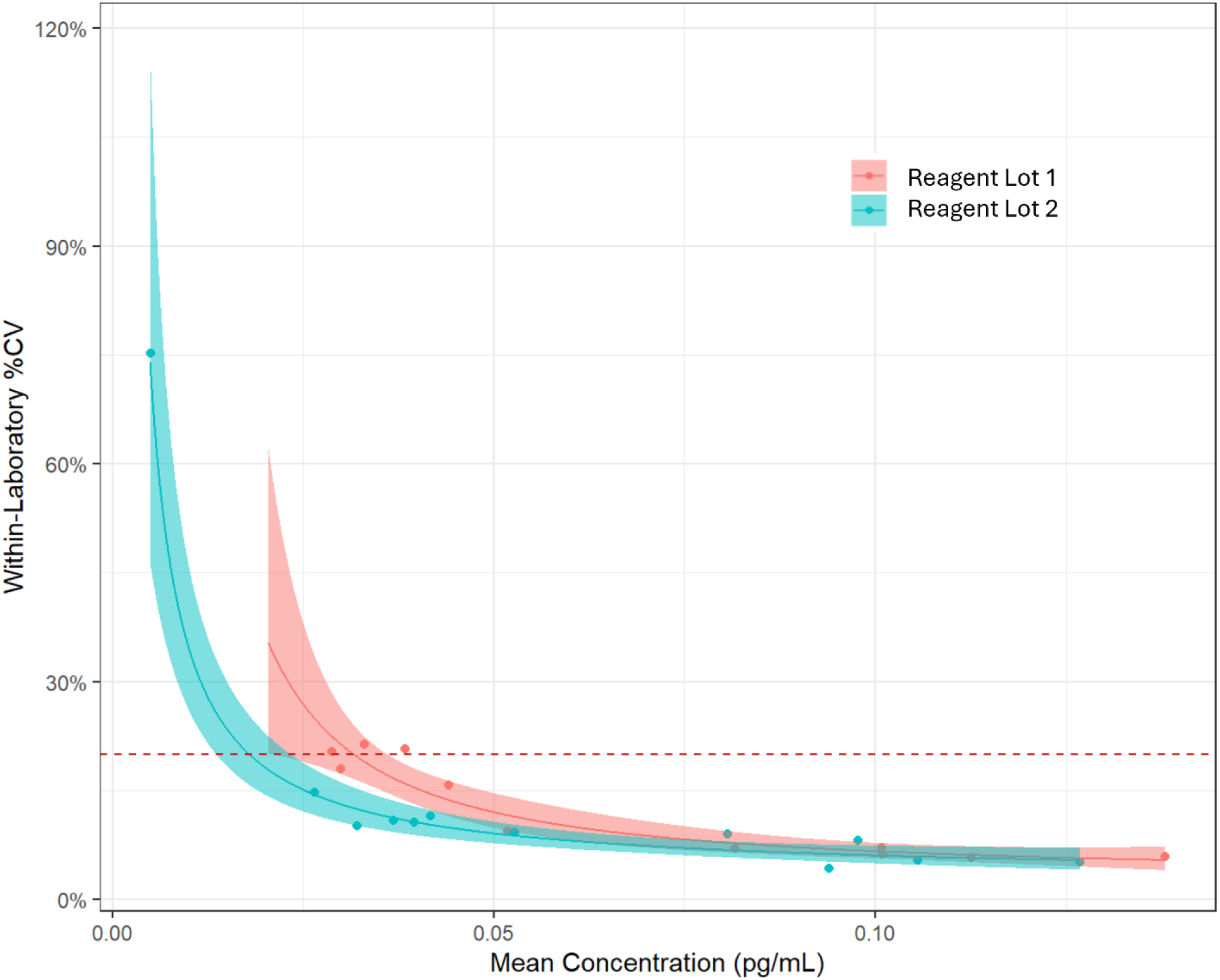
Precision profile and estimated limit of quantitation (LoQ) for the Access p-Tau217 assay across two reagent lots. Within-laboratory precision (%CV) is plotted against mean measured p-Tau217 concentration for low-level EDTA plasma samples. Separate log–log quadratic precision-profile models were fitted for Reagent Lot 1 (red) and Reagent Lot 2 (teal), with shaded bands representing 95% confidence intervals for each fitted curve. Individual observed %CV values for each sample level are shown as points. The dashed horizontal line indicates the 20% total CV criterion used to define the LoQ. Both reagent lots show similar low-end precision characteristics, with LoQ occurring at concentrations well below those observed in research samples, demonstrating the assay’s ability to reliably quantify p-Tau217 at physiologic plasma levels.

#### Linearity

The p-Tau217 assay demonstrated linearity across the full measurement range expected under real-world conditions (0.06 - 1.50 pg/mL) and the full AMR of up to 9.0 pg/mL (Figure 2). Under both scenarios, non-linearity was less than 10% and R² exceeded 0.99. Replicate variability remained low throughout the range, and no evidence of signal saturation or deviation from proportionality was observed at higher concentrations.

**Figure 2.**
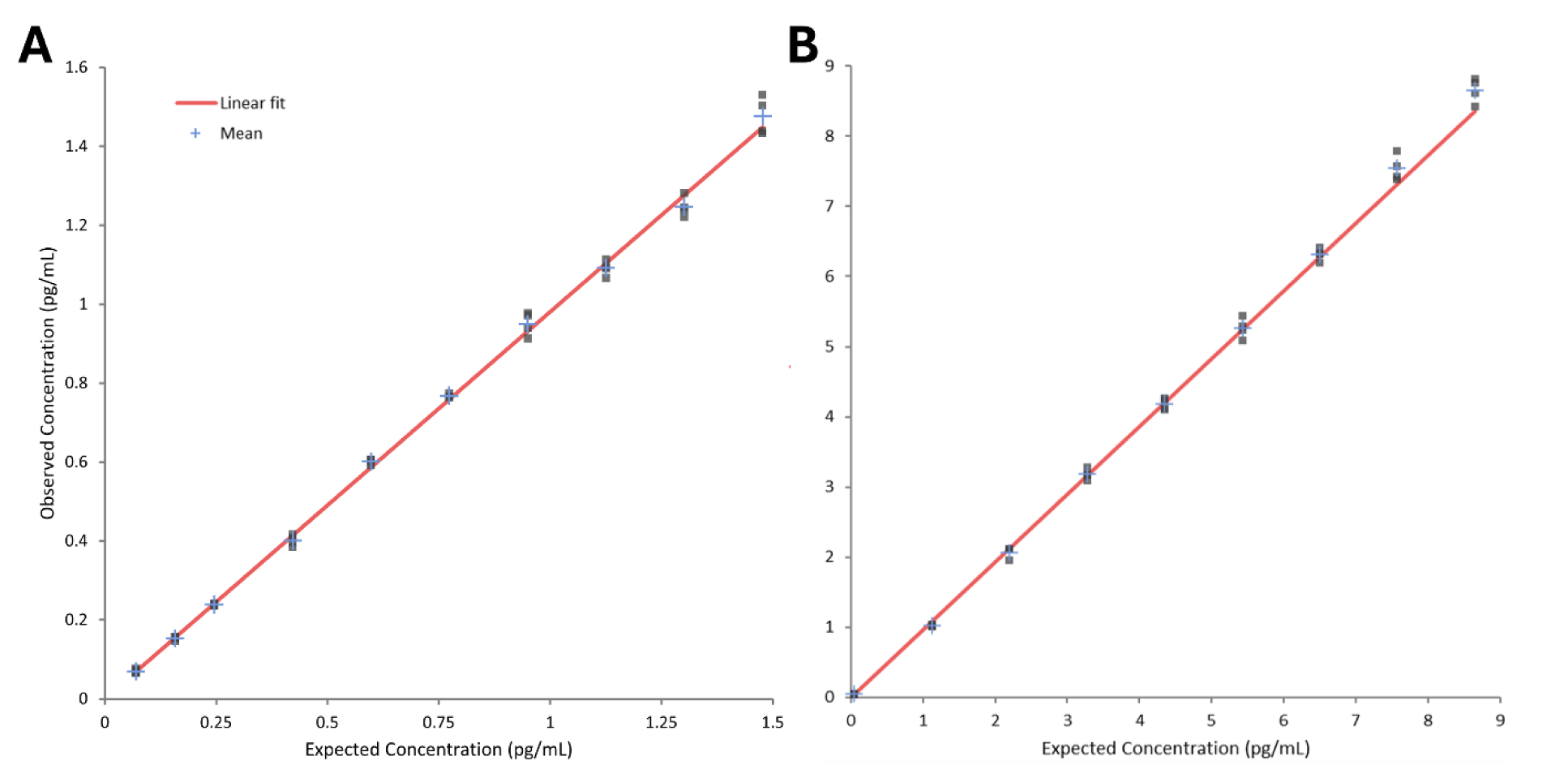
Linearity of the Access p-Tau217. Plasma pools with low and high p-Tau217 concentrations were mixed to generate samples spanning the assay range. Mean observed concentrations (points ± error bars) aligned closely with the linear regression fit (red line) in both the intended-use range (A) and across the full measuring range up to 9 pg/mL (B). Regression models showed R² > 0.99 and non-linearity <10%, demonstrating excellent proportionality across all tested concentrations.

#### Analytical specificity

The p-Tau217 assay demonstrated strong analytical specificity, with low cross reactivity to alternate tau phosphorylation sites. The maximum percentage change in measured p-Tau217 due to spiking with other tau phosphorylation variants was highest for p-tau231, at 1.0%; this was observed for both low (0. 2 pg/mL) and high (0.7 pg/mL) plasma concentrations of p-Tau217 (Table 2). Cross-reactivity was even lower for other tau phosphorylation variants (0 – 0.2% for p-tau181, p-tau205, and p-tau212) and 0% for total tau (Table 2). No significant interference (defined as >10% deviation from baseline) was observed in the presence of more than 60 common endogenous and exogenous substances, including hemoglobin, lipids, bilirubin, and a wide array of medications commonly prescribed in elderly at-risk patients and employed in the treatment of dementia (Supplemental Tables 1 and 2).

**Table 2.**
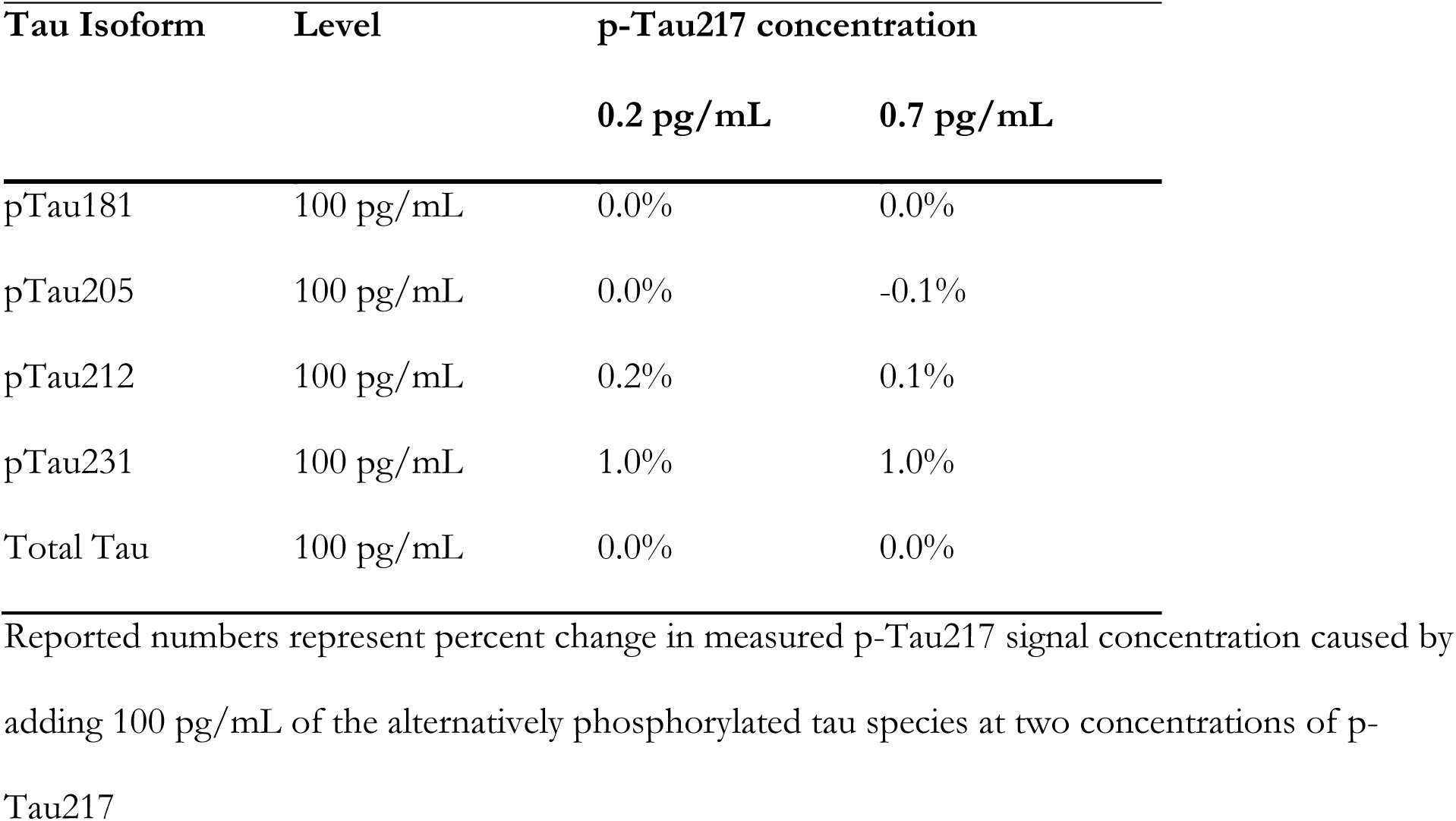
Assay specificity of the p-Tau217 isoform.

| Tau Isoform | Level | p-Tau217 concentration |  |
| --- | --- | --- | --- |
|  |  | 0.2 pg/mL | 0.7 pg/mL |
| pTau181 | 100 pg/mL | 0.0% | 0.0% |
| pTau205 | 100 pg/mL | 0.0% | -0.1% |
| pTau212 | 100 pg/mL | 0.2% | 0.1% |
| pTau231 | 100 pg/mL | 1.0% | 1.0% |
| Total Tau | 100 pg/mL | 0.0% | 0.0% |

Reported numbers represent percent change in measured p-Tau217 signal concentration caused by adding 100 pg/mL of the alternatively phosphorylated tau species at two concentrations of p-Tau217

#### Sample stability

P-Tau217 samples were stable through 48 hours at room temperature (mean difference 4.2%), 48 hours under refrigerated conditions (mean difference 1.1%) and frozen at 30 days (mean difference 0.4%) as seen in Supplemental Table 3. The samples also remained stable over six freeze-thaw cycles (mean difference 5.0%; Supplemental Table 4).

### Discrimination of PET-defined amyloid status

#### Research cohort characteristics

The research cohort comprised 449 participants, including 242 (53.9%) amyloid-negative and 207 (46.1%) amyloid-positive individuals (Table 3). The amyloid-positive group was older than the amyloid-negative group (mean 74.61 vs. 71.80 years). Sex was balanced by amyloid status, with males comprising 48.3% of both amyloid negative and positive participants. Ethnic composition was similar across groups; however, the amyloid-positive group comprised a higher proportion of white participants (93.7% vs. 83.5%) and a lower proportion of black participants (5.3% vs. 13.6%) than the amyloid-negative group. Probable AD diagnosis was almost twice as high in the amyloid-positive group (60.9%) compared to the amyloid-negative group (33.5%). Mean MMSE scores were slightly higher in the amyloid-negative group (26.33) compared to amyloid-positive (24.46). Median plasma p-Tau217 concentrations differed between groups, with higher levels observed in the amyloid-positive group (0.94 pg/mL) compared with the amyloid-negative group (0.35 pg/mL).

**Table 3.** Clinical validation cohort characteristics.

|  | **Overall** | **Amyloid**<br>**Negative** | **Amyloid**<br>**Positive** |
| --- | --- | --- | --- |
| n | 449 | 242 | 207 |
| Age, Mean (SD) | 73.10 (6.44) | 71.80 (6.64) | 74.61 (5.87) |
| Male, No. (%) | 217 (48.3) | 117 (48.3) | 100 (48.3) |
| Ethnicity, No. (%) |  |  |  |
| Hispanic or Latino | 48 (10.7) | 26 (10.7) | 22 (10.6) |
| Not Hispanic or Latino | 394 (87.8) | 215 (88.8) | 179 (86.5) |
| Unknown | 7 (1.6) | 1 (0.4) | 6 (2.9) |
| Race, No. (%) |  |  |  |
| Asian | 5 (1.1) | 3 (1.2) | 2 (1.0) |
| Black | 44 (9.8) | 33 (13.6) | 11 ( 5.3) |
| White | 396 (88.2) | 202 (83.5) | 194 (93.7) |
| Unknown | 4 (0.9) | 4 (1.7) | 0 (0.0) |
| Probable AD Diagnosis, No.<br>(%) | 207 (46.1) | 81 (33.5) | 126 (60.9) |
| MMSE, Mean (SD) | 25.47 (3.03) | 26.33 (2.65) | 24.46 (3.13) |
| p-Tau217 pg/mL, median<br>(IQR) | 0.51 (0.33-0.95) | 0.35 (0.26-0.48) | 0.94 (0.65-1.25) |

Categorical variables are shown as frequencies with percentages in parentheses. Continuous variables are shown as mean with standard deviation (SD) in parentheses or median with interquartile range (IQR) in brackets. AD: Alzheimer's dementia; BEC: Beckman Coulter

#### Biomarker group discrimination

Plasma p-Tau217 demonstrated strong discrimination between amyloid-positive and amyloid-negative participants in the full research cohort (area under the ROC curve [AUC], 0.88 [95% CI 0.85–0.92]). A range of concentration thresholds was evaluated to characterize threshold-dependent behavior of the assay. A single threshold corresponding to the maximum Youden index occurred at 0.580 pg/mL and is shown for reference in Figure 3A. To further explore how overlapping biomarker distributions influence classification characteristics, indeterminate zones ranging from 0% to 30% were applied, generating paired lower and upper concentration thresholds for each scenario. In Figure 3A, these thresholds are displayed along the ROC curve to illustrate how expanding the indeterminate zone alters sensitivity and specificity within the study cohort. Figure 3B shows the empirical distributions of plasma p-Tau217 concentrations for amyloid-negative and amyloid-positive participants with corresponding lower and upper thresholds overlaid, demonstrating how wider indeterminate zones increasingly encompass regions of distributional overlap.

**Figure 3.**
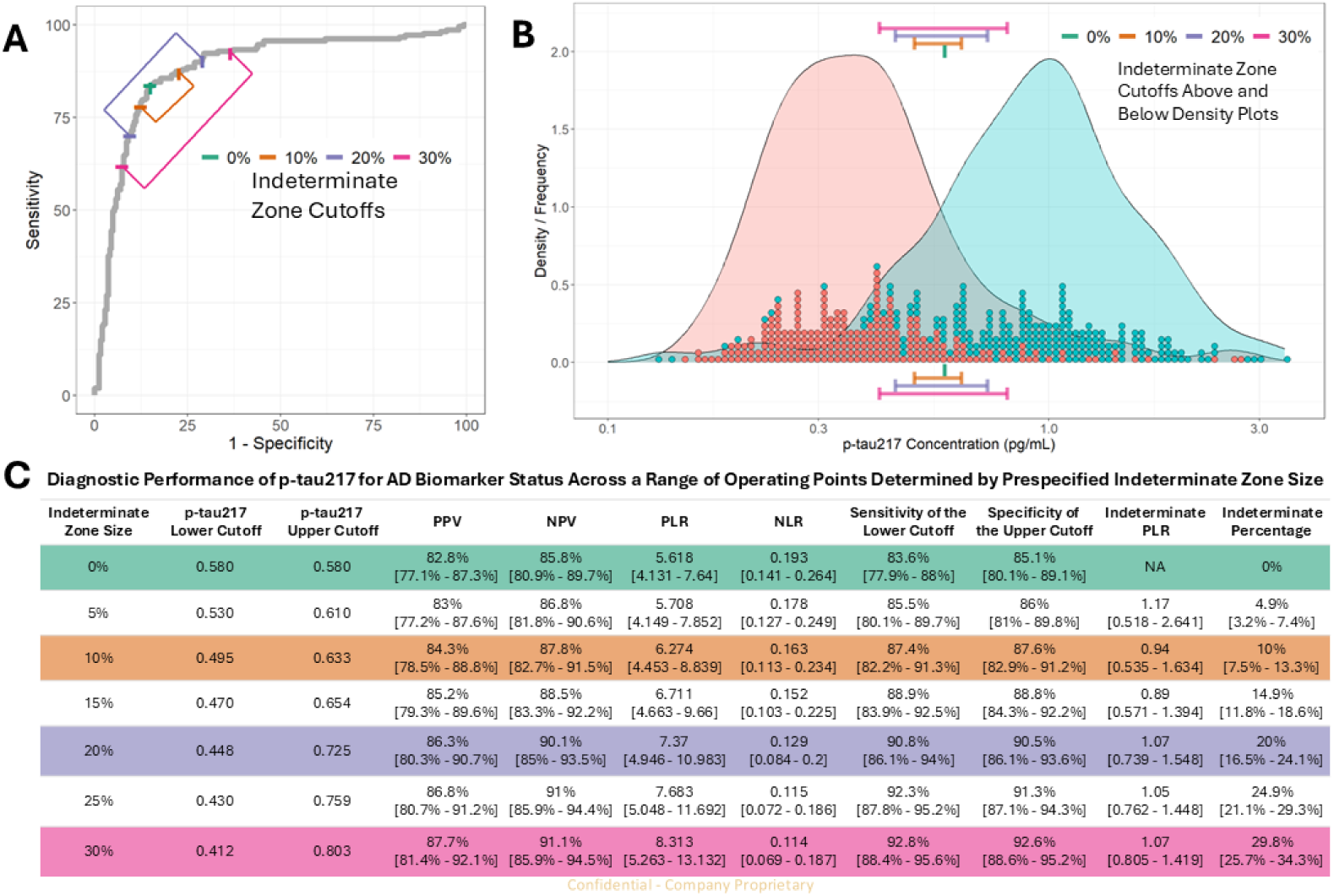
Performance of plasma p-Tau217 with prespecified indeterminate zones for predicting AD amyloid biomarker status. (A) Receiver operating characteristics curve for p-Tau217 assay with superimposed operating points corresponding to increasing indeterminate zone sizes (0%, 10%, 20%, 30%). Each path illustrates how sensitivity and specificity change as progressively wider symmetric indeterminate zones are applied around the classification threshold. (B) Kernel density plots and individual data points showing the distribution of p-Tau217 concentrations among amyloid-negative (red) and amyloid-positive (blue) participants. Horizontal bars indicate the lower and upper p-Tau217 cutoffs for each prespecified indeterminate zone size. The indeterminate zone captures the region where the amyloid-positive and amyloid-negative distributions overlap. (C) Diagnostic performance metrics for p-Tau217 over a range of prespecified indeterminate zone sizes, including positive and negative predictive values (PPV, NPV), positive and negative likelihood ratios (PLR, NLR), sensitivity, specificity, and the proportion of tests designated as indeterminate.

Classification metrics across the full study cohort using prespecified indeterminate zone scenarios are summarized in Figure 3C. With no indeterminate zone, sensitivity was 83.6% and specificity was 85.1%. Application of a 10% indeterminate zone resulted in lower and upper concentration thresholds of 0.495 pg/mL and 0.633 pg/mL, respectively, yielding sensitivity of 87.4%, specificity of 87.6%, a positive predictive value of 84.3%, and a negative predictive value of 87.8%. Application of a 20% indeterminate zone (lower and upper thresholds of 0.448 pg/mL and 0.725 pg/mL) yielded sensitivity of 90.8%, specificity of 90.5%, a positive predictive value of 86.3%, and a negative predictive value of 90.1%. The widest indeterminate zone evaluated (30%) corresponded to thresholds of 0.412 pg/mL and 0.803 pg/mL and resulted in 29.8% indeterminate classifications, with sensitivity of 92.8% and specificity of 92.6%.

## Discussion

In this study, we performed analytical validation of a high-sensitivity plasma p-Tau217 immunoassay and evaluated its ability to discriminate PET-defined amyloid groups in a clinically characterized research cohort. The assay demonstrated robust analytical performance and showed consistent separation between amyloid-positive and amyloid-negative groups. Exploration of multiple concentration thresholds and indeterminate zone widths illustrates how classification performance varies across regions of biomarker overlap. Study findings support the use of the Access p-Tau217 (RUO) immunoassay as a scalable research tool for biological stratification in AD studies, including population enrichment and longitudinal assessment in observational studies and clinical trials.

The Access p-Tau217 (RUO) assay delivered robust analytical performance across the full validation battery. Precision was high (total %CV ≤7%) and detection limits fell well below physiologic p-Tau217 concentrations, indicating that measured values reflect biological signal rather than analytical noise, and linearity across the measuring range showed slopes near unity without signal saturation.

Epitope specificity was strong, with minimal cross-reactivity against other phosphorylated tau isoforms. Interference was negligible across more than 60 endogenous and exogenous substances— including hemolysis, lipids, bilirubin, and medications common in older adults (e.g., statins, cholinesterase inhibitors)—supporting reliable measurement in the heterogeneous plasma samples typical of large-scale biomarker studies.

Although we did not perform a direct head-to-head comparison with other p-Tau217 assays, the discrimination observed in this study (AUC 0.88 [95% CI 0.85–0.92] for PET-defined amyloid status) is consistent with published reports on pTau217 performance.^28,29^ A recent systematic review and meta-analysis identified plasma p-Tau217 as the highest-performing blood-based tau biomarker for biologically defined AD, with a pooled area under the curve of 0.91 (sensitivity 88.1%, specificity 88.7%).^10,11^ Reported discrimination is generally highest in well-characterized specialized and research cohorts—a recent head-to-head comparison, for example, reported areas under the curve of 0.91–0.94 across commercial plasma p-Tau217 immunoassays^30^—and can be modestly lower and more variable in community-recruited, demographically diverse cohorts, such as the cohort studied here.^29,31,32^ Differences in reference-standard methodology (visual amyloid-PET read) and case mix may also contribute. In this context, the discrimination achieved by the Access p-Tau217 (RUO) assay in the diverse, community-based Bio-Hermes-001 cohort is consistent with expectations for this setting; notably, the determinate-sample sensitivity and specificity obtained with a 20% indeterminate zone (both >90%) approached those reported in specialized cohorts. Most recently, Lv and colleagues compared the discriminatory performance of numerous commercially available plasma p-Tau217 assays for PET-defined amyloid positivity, and reported the performance of the Access p-Tau217 (RUO) assay similar or superior to others tested.^33^ Collectively, these findings support the Access p-Tau217 (RUO) assay as a robust biomarker with strong concordance to established amyloid measures and favorable performance across the AD research continuum.

A key contribution of this study is demonstrating that classification behavior can be modulated by varying indeterminate zone width, a generalizable design consideration for blood-based biomarkers measured along a biological continuum. Because AD biology evolves gradually, plasma biomarker concentrations overlap near decision boundaries; expanding the indeterminate zone reduces misclassification by shifting thresholds into regions with greater separation between amyloid-positive and amyloid-negative distributions. Indeterminate zones have precedent in AD biomarker deployment. For example, the FDA-cleared Fujirebio Lumipulse plasma assay incorporates dual cut points for the p-Tau217/Aβ-42 ratio, classifying results as positive, negative, or indeterminate to manage uncertainty near these threshold regions.^15^ Similar dual-threshold approaches in research studies of plasma p-Tau217 benchmarked against CSF or PET reference standards have demonstrated utility when biomarker distributions overlap.^31^ When a 20% indeterminate zone was applied, plasma p-Tau217 classified determinate samples with 90.8% sensitivity (95% CI 86.1– 94.0%) and 90.5% specificity (95% CI 86.1–93.6%) (Figure 3C), achieving the ≥90% sensitivity and specificity criterion recommended by both the Alzheimer’s Association Clinical Practice Guideline and the Global CEO Initiative on Alzheimer’s Disease recommend for a blood-based biomarker to substitute for amyloid PET or cerebrospinal fluid testing in cognitively impaired patients. Notably, the Clinical Practice Guideline cautions that many assays do not reach these thresholds with a single cutoff and explicitly identifies multi-threshold testing as a means of refining diagnostic accuracy— directly motivating the indeterminate-zone approach evaluated here. The 20% deferral fraction observed under this scenario is comparable to that reported for other automated plasma p-Tau217 assays evaluated using two-cutpoint strategies.^34^ As blood-based biomarkers for AD enter routine clinical use, understanding how indeterminate zone design affects classification on automated, high-throughput platforms will be important for optimizing screening workflows and reducing reliance on confirmatory CSF or PET testing.

From a methodological perspective, likelihood ratios provide a prevalence-independent framework for quantifying how threshold strategies influence classification strength. In this study, likelihood ratios varied systematically with indeterminate zone width, illustrating trade-offs between classification confidence and completeness. Narrow zones increase definitive classifications but elevate misclassification risk in overlapping regions, while wider zones reduce misclassification by reserving borderline values for further investigation at the cost more indeterminate results. These trade-offs have informed biomarker implementation across disease areas and will remain relevant as blood-based biomarkers for AD continue to advance through research to clinical pathways.

This study has several limitations that warrant consideration. Although the amyloid PET-characterized cohort examined here, Bio-Hermes-001, was diverse with respect to age, disease stage, and geography of enrollment - biomarker discrimination may differ in settings with lower prevalence of amyloid positivity, such as primary-care or population-based screening settings.^31^ Because the present analysis was restricted to individuals with mild cognitive impairment or mild AD, among whom amyloid positivity was relatively common (46%), the operating characteristics reported here may not extend to cognitively unimpaired, preclinical, or lower-prevalence screening populations, in which both discrimination and predictive values may differ. In addition, the classification thresholds and indeterminate-zone cut points reported here were both derived and evaluated within a single cohort; the resulting operating characteristics are internal estimates that may be optimistic, and independent validation in separate cohorts is required before any threshold is applied more broadly. We were also unable to account for kidney function in this cohort.^35,36^ Because reduced kidney function is associated with higher plasma p-Tau217 concentrations independent of amyloid pathology and can shift optimal classification thresholds, the absence of renal data—particularly in older, community-based populations in which chronic kidney disease is common—is an important limitation. Longitudinal plasma measurements were not available for this analysis, limiting evaluation of within-individual change over time or associations with subsequent clinical trajectories. In addition, while the Bio-Hermes-001 cohort included participants from multiple demographic backgrounds, further evaluation in cohorts with broader racial and ethnic diversity will be important to ensure generalizability of these findings.^32^

In summary, the Access p-Tau217 (RUO) assay demonstrated strong analytical performance and consistent discrimination of PET-defined amyloid status in this research cohort. These findings support its use as a robust, high-throughput tool for biomarker stratification in AD research and may inform the broader translational pathway as plasma p-Tau217 testing is adopted into clinical practice.

## Acknowledgments

We would like to thank the research participants and their families who volunteered time and biospecimens to make this and related research possible. ChatGPT (GPT-5.2, OpenAI) and Claude (Sonnet 5, Anthropic) were used to assist with grammar, readability, and stylistic clarity during manuscript preparation. No AI tools were used for data analysis, interpretation, or scientific decision-making. The authors retain full responsibility for all content.

## Author Contributions

PW and AB contributed equally to study design, data analysis, and manuscript preparation. MH, CC, and JH contributed equally to study conception, supervision, and critical revision of the manuscript. SL contributed to study design, data interpretation, and manuscript revisions. BD, NH, KH, CK, JL, PL, JR, and KB contributed to assay development, analytical validation experiments, and data acquisition. All authors read and approved the final manuscript.

## Statements and Declarations

### Ethical Considerations

The protocol for the Bio-Hermes-001 study was approved by a central institutional review board (Advarra, Reference Number Pro00046018). The study is registered on ClinicalTrials.gov (NCT04733989). Analytical validation studies used commercially available plasma specimens and did not require separate ethical approval.

### Consent to participate

Informed consent was obtained from all study participants.

### Consent for publication

Not applicable.

### Declaration of conflicting interest

Beckman Coulter is the developer and manufacturer of the Access p-Tau217 (RUO) immunoassay studied. PW, AB, MH, CC, BD, NH, KH, CK, JL, PL, JR, and KB were employees of Beckman Coulter at the time of this study. JH and SL are employees of Danaher Diagnostics and hold equity in Danaher Corporation. Both Beckman Coulter and Danaher Diagnostics are wholly owned subsidiaries of Danaher Corporation. HZ has served at scientific advisory boards and/or as a consultant for Abbvie, Acumen, Alamar, Alector, Alzinova, ALZpath, Amylyx, Annexon, Apellis, Artery Therapeutics, AZTherapies, Bioventix, Cognitact, Cognito Therapeutics, CogRx, Denali, Eisai, Enigma, Johnson & Johnson, LabCorp, Merck Sharp & Dohme, Merry Life, Nervgen, Neurocode, New Amsterdam, Novo Nordisk, Optoceutics, Passage Bio, Pinteon Therapeutics, Prothena, Quanterix, Red Abbey Labs, reMYND, Roche, Samumed, ScandiBio Therapeutics AB, Siemens Healthineers, Triplet Therapeutics, and Wave, has given lectures sponsored by Alzecure, BioArctic, Biogen, Cellectricon, Fujirebio, LabCorp, Lilly, Novo Nordisk, Oy Medix Biochemica AB, Roche, and WebMD, is a co-founder of Brain Biomarker Solutions in Gothenburg AB (BBS), which is a part of the GU Ventures Incubator Program, and is a shareholder of CERimmune Therapeutics (outside submitted work).

### Funding statement

This study was performed without external funding.

### Data availability

The datasets generated and analyzed during the current study are not publicly available due to restrictions associated with source cohort agreements.

