## Supplemental data for "Analytical validation and amyloid-status discrimination of a high-throughput, research-use-only plasma p-Tau217 immunoassay"

**Supplemental Material**

**Supplemental Methods. Specimen sources for analytical characterization**

Plasma specimens used for each analytical characterization study were obtained from the following commercial biospecimen vendors:

**Precision:** BioIVT, Dx Biosamples, iSpecimen, SignalChem, and Grifols.

**Analytical sensitivity:** BioIVT.

**Linearity:** BioIVT.

**Analytical specificity:** BioIVT.

Selected samples were supplemented with recombinant p-Tau217 (Tau-441 phosphorylated by DYRK1A) to achieve target concentrations.

**Supplemental Table 1. Endogenous interference**

| **Interferent** | **Interferent Level** | **p-Tau217 concentration** | |
|---|---|---|---|
| | | **0.2 pg/mL** | **0.7 pg/mL** |
| Hemoglobin | 3.3 mg/mL | -2.3% | N/A |
| Conjugated Bilirubin | 40 mg/dL | 4.0% | 0.3% |
| Unconjugated Bilirubin | 40 mg/dL | -1.9% | -3.2% |
| Triglycerides | 1350 mg/dL | 0.7% | 8.6% |
| Human Serum Albumin | 140 mg/mL | -6.4% | -9.5% |
| Immunoglobulin (IgG) | 5 mg/mL | -5.1% | -2.7% |
| Rheumatoid Factor | 30 IU/mL | 7.6% | N/A |
| Human Anti-Mouse Antibody (HAMA) | 1000 ng/mL | -3.4% | -1.1% |



Confidential - Company Proprietary

Reported numbers represent percent change in measured p-Tau217 signal concentration at two plasma concentrations caused by adding specified concentrations of each substance.

**Supplemental Table 2. Exogenous interference**

| Interferent | Interferent Level | p-Tau217 concentration | |
|---|---|---|---|
| | | **0.2 pg/mL** | **0.7 pg/mL** |
| Acetaminophen | 20 mg/dL | 3.4% | -2.0% |
| Acetazolamide | 5.7 mg/dL | 4.8% | 0.6% |
| Acetylcysteine | 15.0 mg/dL | 5.0% | -4.7% |
| Acetylsalicyclic Acid | 100 mg/dL | 0.3% | -0.5% |
| Ampicillin | 100 mg/dL | 1.6% | 0.5% |
| Aripiprazole | 45 mg/L | -2.3% | 0.6% |
| Ascorbic Acid | 60.23 mg/dL | -4.2% | -6.0% |
| Atorvastatin | 0.075 mg/dL | 0.3% | -1.0% |
| Biotin | 3500 ng/mL | -0.4% | 2.2% |
| Bisacodyl | 0.009 mg/mL | 0.4% | -0.2% |
| Caffeine | 556 µmol/L | -5.2% | -2.2% |
| Cefoxitin | 660 mg/dL | -1.7% | -0.6% |
| Chloramphenicol | 241 µmol/L | -1.0% | -1.5% |
| Chlorothiazide | 2.7 mg/dL | 1.0% | 5.8% |
| Clopidogrel | 0.045 mg/mL | -4.2% | -0.2% |
| Cyclosporine | 0.5 mg/dL | 0.9% | -0.8% |


Confidential - Company Proprietary

| | | | |
|---|---|---|---|
| Desipramine HCL | 0.204 mg/dL | -1.3% | -1.3% |
| Diazepam | 3 mg/dL | -0.9% | -0.1% |
| Digoxin | 0.004 mg/dL | 5.1% | 0.9% |
| Dobesilate Calcium | 6 mg/dL | 3.3% | -6.0% |
| Donepezil (Aricept) | 30 mg/L | -6.1% | 0.3% |
| Doxycycline | 1.8 mg/dL | -1.5% | -1.5% |
| Escitalopram | 0.0192 mg/dL | 2.5% | 2.2% |
| Esomeprazole | 0.69 mg/dL | 1.2% | 1.5% |
| Estradiol | 0.00000075 mg/dL | -2.8% | 1.7% |
| Famotidine | 0.244 mg/dL | -3.3% | -0.2% |
| Furosemide | 1.59 mg/dL | -2.7% | -5.4% |
| Gabapentin | 2.67 mg/dL | -7.4% | 0.5% |
| Galantamine (Reminyl) | 250 mg/L | 2.6% | -1.0% |
| Heparin | 330 units/dL | -6.1% | -1.8% |
| Hydralazine HCL | 1.44 mg/dL | -2.1% | 1.1% |
| Hydrochlorothiazide | 0.113 mg/dL | 0.7% | 5.0% |
| Ibuprofen | 50 mg/dL | -2.5% | 0.8% |
| Lactitol | 12 mg/mL | 2.3% | 4.4% |
| Levodopa | 2 mg/dL | -6.0% | 2.2% |
| Levothyroxine | 0.0429 mg/dL | -4.0% | -0.1% |
| Lisinopril | 0.0246 mg/dL | -2.7% | -0.1% |
| Loperamide HCl | 0.0096 mg/mL | 2.0% | -3.8% |
| Losartan | 0.06 mg/mL | 2.8% | 2.6% |
| Memantine (Namenda) | 250 mg/L | -2.9% | -1.6% |



Confidential - Company Proprietary

| | | | |
|---|---|---|---|
| Metformin | 1.2 mg/dL | -0.6% | 0.1% |
| Methyldopa | 2.25 mg/dL | -6.6% | 0.8% |
| Minocycline | 0.12 mg/mL | -9.7% | 3.1% |
| Mirtazapine | 0.027 mg/mL | -1.0% | 2.3% |
| Nifedipine | 5.4 mg/dL | -2.2% | -1.0% |
| Phenylbutazone | 32.1 mg/dL | -8.7% | -2.1% |
| Prednisone | 0.0099 mg/dL | -1.7% | 2.3% |
| Rifampicin | 6 mg/dL | 6.6% | 1.4% |
| Rivastigmine (Exelon) | 45 mg/L | -2.8% | -2.3% |
| Sacubitril Calcium Salt (Entresto) | 0.915 mg/dL | -2.9% | -7.0% |
| Simvastatin | 0.168 mg/dL | -7.2% | 0.8% |
| Spironolactone | 0.0555 mg/dL | 0.5% | 3.4% |
| Sulfasalazine | 7.5 mg/dL | 1.9% | -1.9% |
| Theophylline | 10 mg/dL | -2.1% | 2.6% |
| Tramadol | 0.314 mg/dL | 1.3% | -0.5% |
| Valsartan | 1.17 mg/dL | -9.3% | -0.4% |
| Venlafaxine HCl | 0.135 mg/mL | 5.1% | -4.1% |
| Zaleplon | 0.012 mg/mL | -2.5% | -2.5% |

Reported numbers represent percent change in measured p-Tau217 signal concentration at two plasma concentrations caused by adding specified concentrations of each substance.

**Supplemental Table 3. Sample stability over time**



Confidential - Company Proprietary

| Time | % Difference from Baseline | | |
|---|---|---|---|
| | Room Temp | Refrigerated | Frozen at -20°C |
| 6 hours | -2.1% | - | - |
| 24 hours | 1.3% | 0.1% | - |
| 48 hours | -4.2% | -1.1% | - |
| 30 days | - | - | -0.4% |

**Supplemental Table 4. Sample stability over freeze/thaw cycles**

| # Freeze/thaws | % Difference from Baseline |
|---|---|
| 2 | 0.9% |
| 3 | -2.4% |
| 4 | 0.5% |
| 5 | -3.3% |
| 6 | -5.0% |



Confidential - Company Proprietary

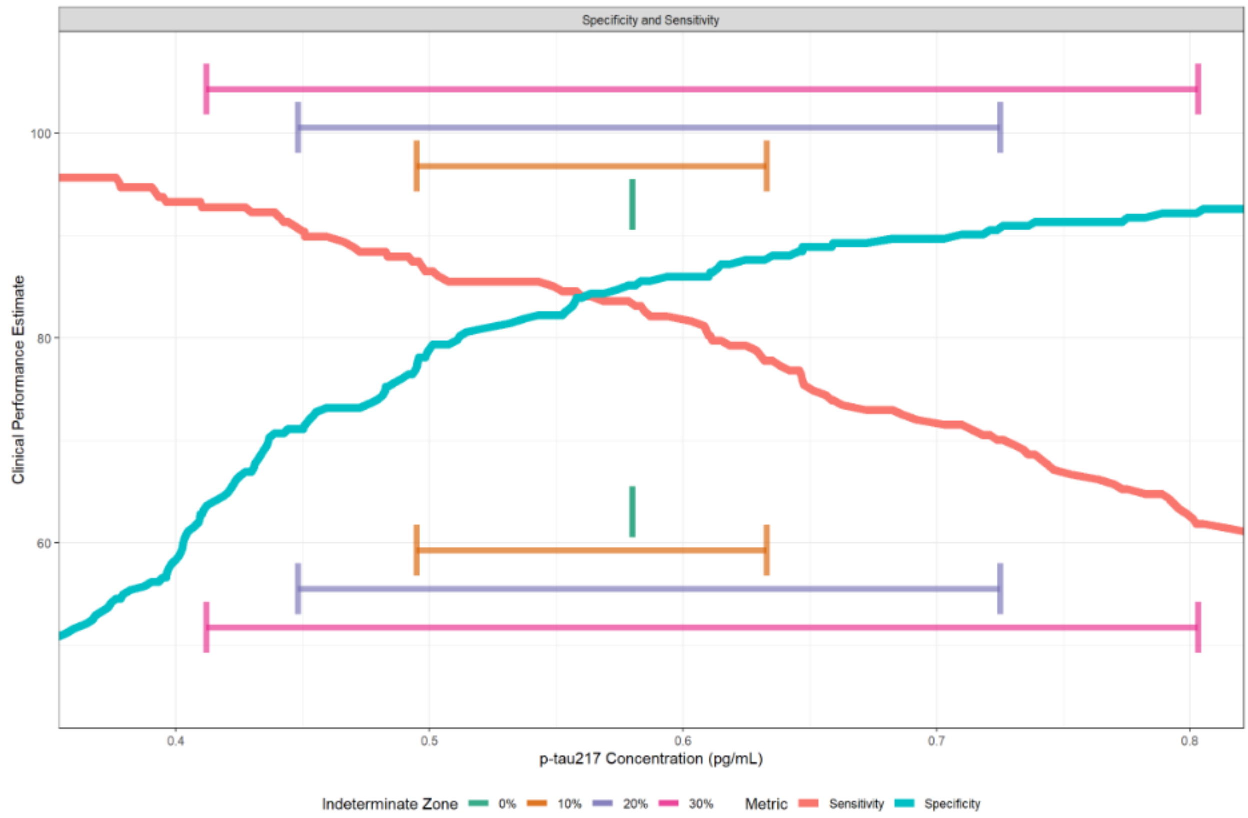


**Supplemental Figure 1. Sensitivity and specificity performance curves across the indeterminate-zone threshold range**

Empirical sensitivity of the lower cutoff (red) and specificity of the upper cutoff (teal) curves for plasma p-Tau217 plotted against concentration thresholds spanning the range explored across the 0–30% indeterminate-zone scenarios shown in main Figure 3. Horizontal range bars indicate the lower and upper concentration cutoffs for each indeterminate-zone width (0%, 10%, 20%, 30%); the same cutoffs are overlaid at the upper and lower extents of the plot for visual alignment with the sensitivity curve (top) and the specificity curve (bottom).



Confidential - Company Proprietary

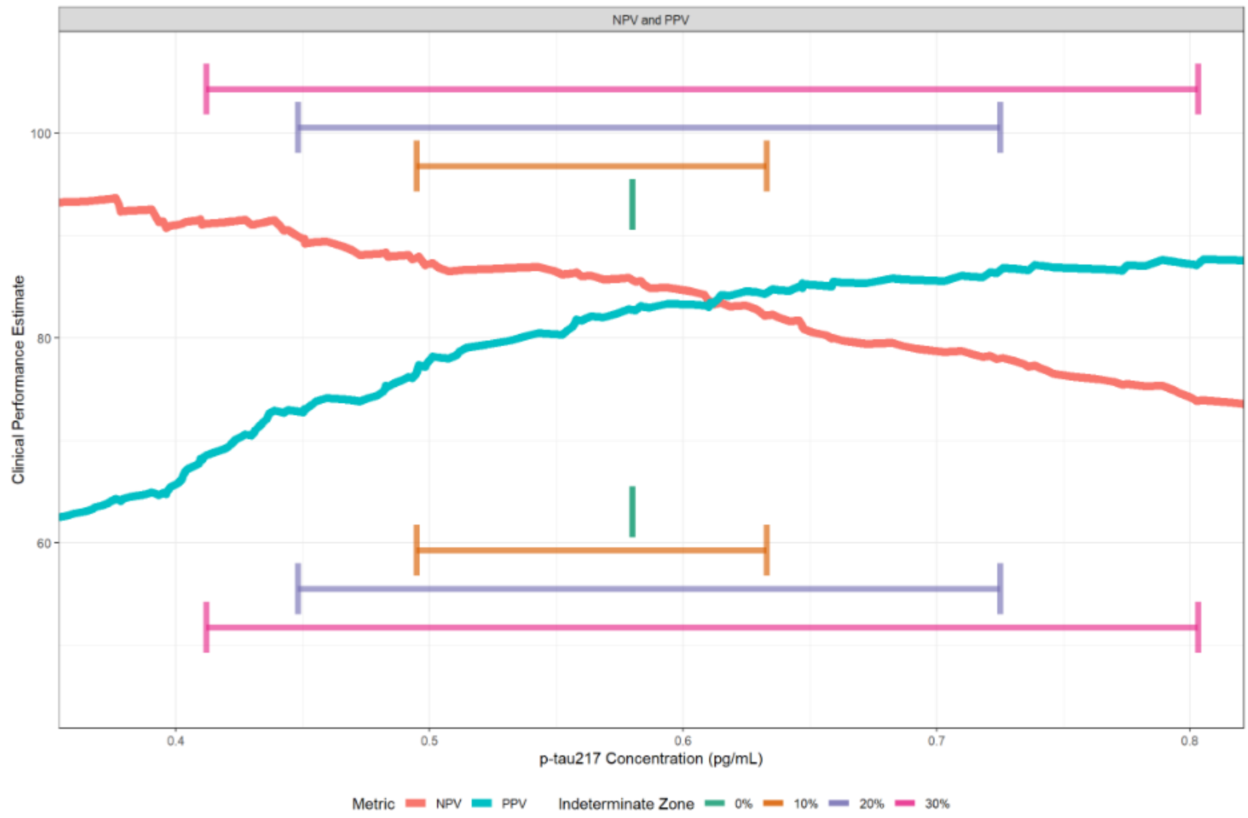


**Supplemental Figure 2. Positive and negative predictive value performance curves across the indeterminate-zone threshold range**

Empirical positive predictive value (PPV; teal) and negative predictive value (NPV; red) curves for plasma p-Tau217 plotted against concentration thresholds spanning the range explored across the 0–30% indeterminate-zone scenarios shown in main Figure 3. Horizontal range bars indicate the lower and upper concentration cutoffs for each indeterminate-zone width (0%, 10%, 20%, 30%); the same cutoffs are overlaid at the upper and lower extents of the plot for visual alignment with each metric curve.



Confidential - Company Proprietary

**Supplemental Table 5. Indeterminate zone size at different p-Tau217 cutoff locations**

| Indeterminate Zone at different cutoff combinations. Performance reported as Sensitivity of the Lower Cutoff and Specificity of the Upper Cutoff | | Upper Cutoff | | | | | | |
|---|---|---|---|---|---|---|---|---|
| | | Cutoff Location: 0.412 Spec: 63.6 | Cutoff Location: 0.448 Spec: 71.1 | Cutoff Location: 0.495 Spec: 77.3 | Cutoff Location: 0.580 Spec: 85.1 | Cutoff Location: 0.633 Spec: 87.6 | Cutoff Location: 0.725 Spec: 90.5 | Cutoff Location: 0.803 Spec: 92.6 |
| Lower Cutoff | Cutoff Location: 0.412 Sens: 92.8 | 0 | 4.9 | 9.8 | 15.8 | 19.8 | 24.9 | 29.8 |
| | Cutoff Location: 0.448 Sens: 90.8 | | 0 | 4.9 | 10.9 | 14.9 | 20.0 | 24.9 |
| | Cutoff Location: 0.495 Sens: 87.4 | | | 0 | 6.0 | 10.0 | 15.1 | 20.0 |
| | Cutoff Location: 0.580 Sens: 83.6 | | | | 0 | 4.0 | 9.1 | 14.0 |



Confidential - Company Proprietary

| | | | | | | | | |
|---|---|---|---|---|---|---|---|---|
| | Cutoff<br>Location: 0.633<br>Sens: 77.8 | | | | | 0 | 5.1 | 10.0 |
| | Cutoff<br>Location: 0.725<br>Sens: 70.0 | | | | | | 0 | 4.9 |
| | Cutoff<br>Location: 0.803<br>Sens: 61.8 | | | | | | | 0 |



Confidential - Company Proprietary
